# Uncovering the genetic underpinnings for different psychiatric disorder combinations

**DOI:** 10.1101/2024.08.29.24312761

**Authors:** Liangying Yin, Menghui Liu, Yujia Shi, Ruoyu Zhang, Simon Sai-Yu Lui, Hon-Cheong So

**Affiliations:** School of Biomedical Sciences, The Chinese University of Hong Kong, Shatin, Hong Kong; Department of Psychiatry, School of Clinical Medicine, The University of Hong Kong, Hong Kong SAR, China; KIZ-CUHK Joint Laboratory of Bioresources and Molecular Research of Common Diseases, Kunming Institute of Zoology and The Chinese University of Hong Kong, China; Department of Psychiatry, The Chinese University of Hong Kong, Hong Kong; CUHK Shenzhen Research Institute, Shenzhen, China; Margaret K.L. Cheung Research Centre for Management of Parkinsonism, The Chinese University of Hong Kong, Shatin, Hong Kong; Brain and Mind Institute, The Chinese University of Hong Kong, Hong Kong SAR, China; Hong Kong Branch of the Chinese Academy of Sciences Center for Excellence in Animal Evolution and Genetics, The Chinese University of Hong Kong, Hong Kong SAR, China

## Abstract

Psychiatric disorders are highly heterogeneous. Clinically, it is meaningful to distinguish psychiatric disorders by the presence or absence of a specific comorbid condition. In this study, we employed a recently developed algorithm (CombGWAS) to decipher the genetic basis of psychiatric disorder combinations using GWAS summary statistics. We focused on comorbidities and combinations of diseases, such as schizophrenia(SCZ) with and without depression, which can be considered as two “subtypes” of SCZ. We also studied psychiatric disorders comorbid with obesity as disease subtypes.

Our study identified genetic signatures that distinguish different “subtypes” and revealed the genetic architecture underlying 16 psychiatric disorder combinations. We discovered both shared and unique susceptibility genes/variants across different psychiatric disorder “subtypes”.

Notably, despite high genetic correlations between subtypes, most subtype pairs exhibited distinct genetic correlations with the same cardiovascular disease (CVD). Some pairs even displayed opposite genetic correlations, especially those involving obesity. For instance, the genetic correlation (rg) between SCZ with obesity and type 2 diabetes(T2DM) was 0.248(p=4.42E-28), while the rg between SCZ without obesity and T2DM was -0.154(p=6.79E-12).

Many psychiatric disease combinations studied were causally associated with increased CVD risks. Moreover, comorbid psychiatric disorders often showed different causal relationships with CVD compared to single psychiatric disorders. This study represents a significant step toward understanding the genetic heterogeneity underlying psychiatric disorder comorbidities, providing new insights into their pathophysiology.

## Introduction

Psychiatric disorders constitute significant health and societal burdens on a global scale, consistently ranked among the most debilitating medical conditions across diverse age groups^1^. While diagnostic classification systems define disorders based on symptomatology, psychiatric comorbidity is common, with patients frequently presenting with multiple concurrent conditions. For example, a considerable proportion of schizophrenia patients have comorbid depression, obsessive compulsive disorder (OCD) and other psychiatric disorders^2^. Similarly, around 75% of patients with depression have comorbid anxiety disorders.

This comorbidity highlights the vast heterogeneity in psychiatry, as subtypes of a primary disorder with different psychiatric comorbidities may have distinct neurobiological underpinnings. Clarifying the genetic basis for these different psychiatric co-morbidities can unveil biological mechanisms and promote individualized care.

Genome-wide association studies (GWAS) have achieved success in revealing the genetic basis of complex disorders^3,4^, including psychiatric disorders. However, prior research on GWAS mainly focused on exploring the genetic basis of a *single* psychiatric entity, and largely ignored psychiatric comorbidity.

Our team has recently developed an innovative statistical framework, CombGWAS^5^, to uncover the genetic architecture of disease combinations. In this study, we applied CombGWAS to decipher the genetic signatures distinguishing 16 psychiatric disorder combinations, such as schizophrenia and major depressive disorder (MDD) with and without various comorbidities. We also considered psychiatric disorders with/without comorbid obesity as disease subtypes, highlighting the intricate relationships between these conditions and their implications for cardiovascular health.^5^

Unlike other conventional methods such as polygenic risk scores (PRS) ^6^ or LD score regression (LDSR) ^7^ that only reveal *overall* genetic overlap, our approach can identify specific susceptibility variants responsible for the presence or absence of a particular comorbidity. This provides deeper insights into the complex genetic underpinnings of psychiatric disorder heterogeneity.

Our method can also estimate the effect size (e.g. odds ratio) of the variants contributing to the comorbidities, which cannot be achieved by other existing approaches. Furthermore, we employed additional analyses, including genetic correlation, Mendelian randomization (MR), and functional enrichment, to comprehensively characterize the shared and unique genetic architectures across psychiatric subtypes and their relationships with cardiovascular diseases.

57685

We summarize the key contributions and novelties of this study below□

1. We applied a novel statistical framework, CombGWAS, to decipher the genetic signatures that distinguish 16 psychiatric disorder combinations, including subtypes with and without specific comorbidities. We identified numerous susceptibility genes/variants across different psychiatric disease subtypes, providing insights into the genetic heterogeneity underlying these conditions.
2. We investigated MDD and SCZ with/without obesity as disease subtypes and revealed their genetic underpinnings. To our knowledge, this is the first GWAS-based study to uncover genetic variants underlying these comorbid conditions.
3. We revealed distinct genetic correlations between psychiatric disorder subtypes and cardiovascular diseases, shedding light on the complex links between psychiatric and cardiometabolic traits. For example, we uncovered that MDD and SCZ with or without obesity exhibit very different, and often opposite genetic correlations with cardiometabolic disorders.
4. We found that comorbid psychiatric disorders often have more profound causal effects on cardiovascular risks compared to single psychiatric disorders, but the causal effect estimates also vary widely across different psychiatric subtypes. For example, obese and non-obese MDD/SCZ showed opposite causal effects on risks of T2DM.
5. We provided comprehensive genetic and biological insights (through pathway, cell-type, and drug enrichment analyses) that may inform the development of personalized treatment and prevention strategies for psychiatric disorder combinations.

Overall, our work represents a significant advancement in understanding the complex genetic underpinnings of psychiatric disorder heterogeneity and comorbidities, which is crucial for improving clinical management and patient outcomes in the long term.

## Method

In this study, we employed CombGWAS to estimate the genetic architecture of the combinations of psychiatric disorders. CombGWAS is a statistical framework specially designed to mimic case-control GWAS by using summary statistics of the corresponding diseases only. The mathematical formulation is as follows.

Consider *s* ∼ *bin*(*n* = 2, *q*) as a random biallelic SNP with *q* denoting the effect allele frequency. We considered two binary psychiatric-related traits *P*_l_ and *P*_2_, and generated a multivariate linear model on N subjects as follows:

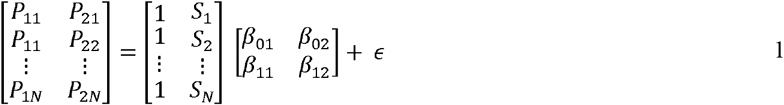

The left-hand side is a *N* × 2 phenotype matrix, the matrices on the right-hand side are respectively an *N* × 2 SNP matrix, 2 × 2 association estimates matrix, and an *N* × 2 error matrix. The error matrix (ϵ ∼*N*(0, ∑)) is assumed to follow a multivariate normal distribution.

The psychiatric comorbidity of two disorders (*P*_*1*_ and *P*_*2*_) can be defined as a function of these two traits:

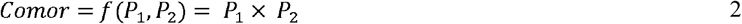

Similarly, the ‘single’ psychiatric disorder (with the particular comorbid disorder excluded) can be defined as follows:

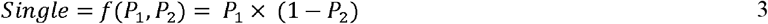

Our goal here is to infer the association estimates for genome-wide SNPs for the target traits (i.e., the comorbid psychiatric disorders or the single psychiatric disorder without the specific comorbid disorder)

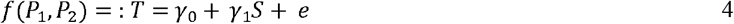

Here, *e* is assumed to follow a normal distribution with zero mean. According to Yin et al.^5^, the association estimates for the combined psychiatric disorders can be estimated as follows:

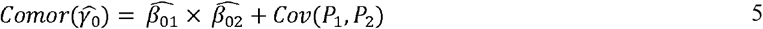

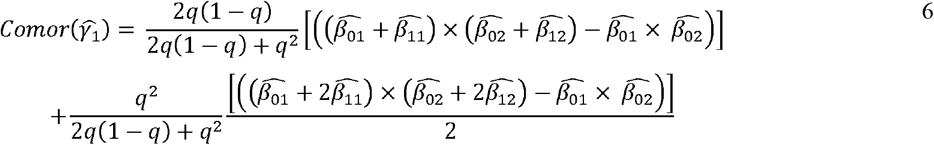

Here, *Cov*(*P*_l_,*P*_2_) denotes the covariance between the two binary traits. Along the same line, the association estimates for the single psychiatric disorder without the particular comorbid disorder can be computed as follows:

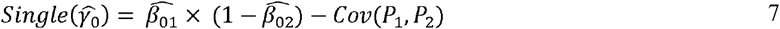

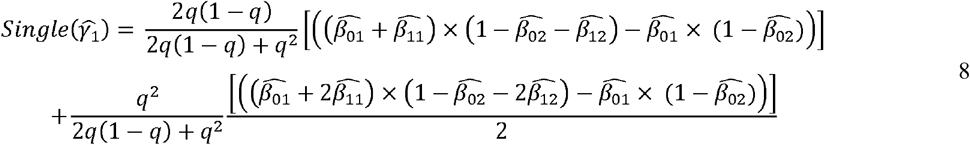

It is noteworthy that the estimates used in the above equations are presumably derived from a linear regression model. However, for binary traits, the association estimates are usually calculated from logistic regression models. Therefore, we need to transform the estimates (odds ratios) derived from logistic regression to that of linear regression, using the following equation:

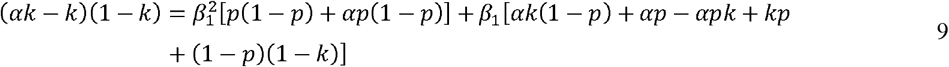

where *p* indicates the effect allele frequency of the SNP under study *S, k* represents the proportion of cases and β_1_ represents the coefficient under a linear model, *α* defines the odds ratio of SNP *S* regressed on the same binary trait. The variances for 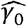 and 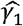 for the target disease combination can be estimated by the delta method, which employs first-order Taylor expansion to estimate the variance of a function. Also note that our proposed analytic framework assumes the absence of significant interactions between the comorbid trait(s) and the genetic variants under study, as explained elsewhere^5^. In our previous study ^5^, we have verified the validity of CombGWAS both using extensive simulations and applications to the UK-Biobank data. Briefly, we re-performed a GWAS in UKBB using the actual phenotype data of disease combinations as the outcome, and found that the results were highly similar to those computed from CombGWAS.

### Enrichment analyses

We utilized several methods to clarify the putative functional and biological mechanisms of the identified susceptibility variants for the “psychiatric phenotypes/subtypes” of interest (i.e., the comorbid psychiatric disorders, and the ‘single’ psychiatric disorders without the particular comorbid disorder). First, the MAGMA tool within FUMA ^8,9^ was used to map the identified genetic variants to their corresponding genes. This established the gene-level associations with the psychiatric phenotypes/subtypes. The definitions for independent significant SNPs and genomic risk loci in our study were also based on the criteria established by FUMA (see https://fuma.ctglab.nl/tutorial#riskloci).

Second, we performed tissue specificity analysis to examine if the susceptibility genes were enriched in specific tissues. Furthermore, we carried out a cell type enrichment analysis to uncover specific cell types or cell populations that may play crucial roles in the etiology of the subtypes. We also conducted pathway analysis using the program “ConsensusPathDB”^10,11^. Moreover, we performed drug enrichment analyses on the identified susceptibility gene sets using “enrichr” ^12,13^. This analysis identified potential therapeutic agents or drug targets that may be relevant to the treatment or management of the investigated psychiatric conditions.

### Genetic correlation among psychiatric disease subtypes and cardiovascular diseases

We employed LD score regression (LDSR) ^7^ to calculate the genetic correlation between different psychiatric entities to see if they represent genetically distinct disease subtypes with unique genetic architectures. Furthermore, we explored the genetic correlations between different psychiatric disorder subtypes and CVDs, including coronary artery disease (CAD), type 2 diabetes (T2DM), and stroke, highlighting possible links between these conditions.

### Mendelian Randomization (MR) analysis

Mendelian Randomization (MR) is a method which leverages genetic variants as ‘instruments’ to represent the exposure for inferring causal relationships between risk factors and outcomes. We performed MR to estimate the causal effects of the studied psychiatric entities on various cardiovascular diseases. A two-sample MR design was employed, using both inverse-variance weighted’ (MR-IVW) ^14^ and Egger regression (MR-Egger) ^15^ approaches. The number of genetic variants included in the MR analysis may influence the causal estimates. To ensure the robustness of our findings, we performed MR at multiple *r*^2^ thresholds (0.001, 0.01, 0.05, and 0.1), taking SNP correlations into account.

### Psychiatric disorders studied

We applied CombGWAS to examine 7 different psychiatric disorders and obesity, i.e., Alzheimer’s disease (AD), attention deficit hyperactivity disorder (ADHD), autism spectrum disorder (ASD), anxiety disorders, insomnia, major depressive disorder (MDD), obesity, and schizophrenia (SCZ). The datasets used are shown in Supplementary Table S1. Using CombGWAS, we estimated the effect sizes (odds ratios (ORs)), standard errors (SEs) and corresponding p-values for each disease combination for genome-wide SNPs.

We analyzed the genetic basis for 16 (or 8 pairs of) psychiatric entities, with a primary focus on MDD and SCZ. The psychiatric entities include ADHD with and without comorbid ASD, MDD with and without AD, MDD with and without Anxiety Disorders, MDD with and without ASD, MDD with and without Insomnia, MDD with and without Obesity, MDD with and without SCZ, SCZ with and without Obesity.

## Results

### Overview

Table 1 demonstrates the independent significant SNPs (p-value<5e-08, r^2^<0.6) and genomic risk loci for the 8 comorbid psychiatric disorders and 8 single psychiatric entities (single psychiatric disorder without a particular comorbid disorder). Our analysis revealed a total of 954 independent significant SNPs, mapped to 325 genomic risk loci, for the 8 comorbid psychiatric disorders (Figure 1, Table 1 and Table S2). In addition, we identified 934 independent significant SNPs mapped to 328 genomic risk loci for the 8 single psychiatric entities (see Figure 1, Table 1 and Supplementary Table S2).

**Table 1.** Number of identified independent significant SNPs and genomic risk loci for different combinations of psychiatric disorders.

| Disease subtypes | No. of ind. sig. SNPs | No. risk loci | Disease subtypes | No. of ind. sig. SNPs | No. risk loci |
| --- | --- | --- | --- | --- | --- |
| ADHD with ASD | 20 | 19 | ADHD without ASD | 23 | 12 |
| MDD with AD | 148 | 37 | MDD without AD | 153 | 53 |
| MDD with Anxiety | 209 | 73 | MDD without Anxiety | 139 | 52 |
| MDD with ASD | 3 | 3 | MDD without ASD | 149 | 51 |
| MDD with Insomnia | 111 | 48 | MDD without Insomnia | 111 | 39 |
| MDD with Obesity | 50 | 21 | MDD without Obesity | 44 | 21 |
| MDD with SCZ | 330 | 95 | MDD without SCZ | 146 | 52 |
| SCZ with Obesity | 83 | 29 | SCZ without Obesity | 169 | 48 |

**Figure 1.**
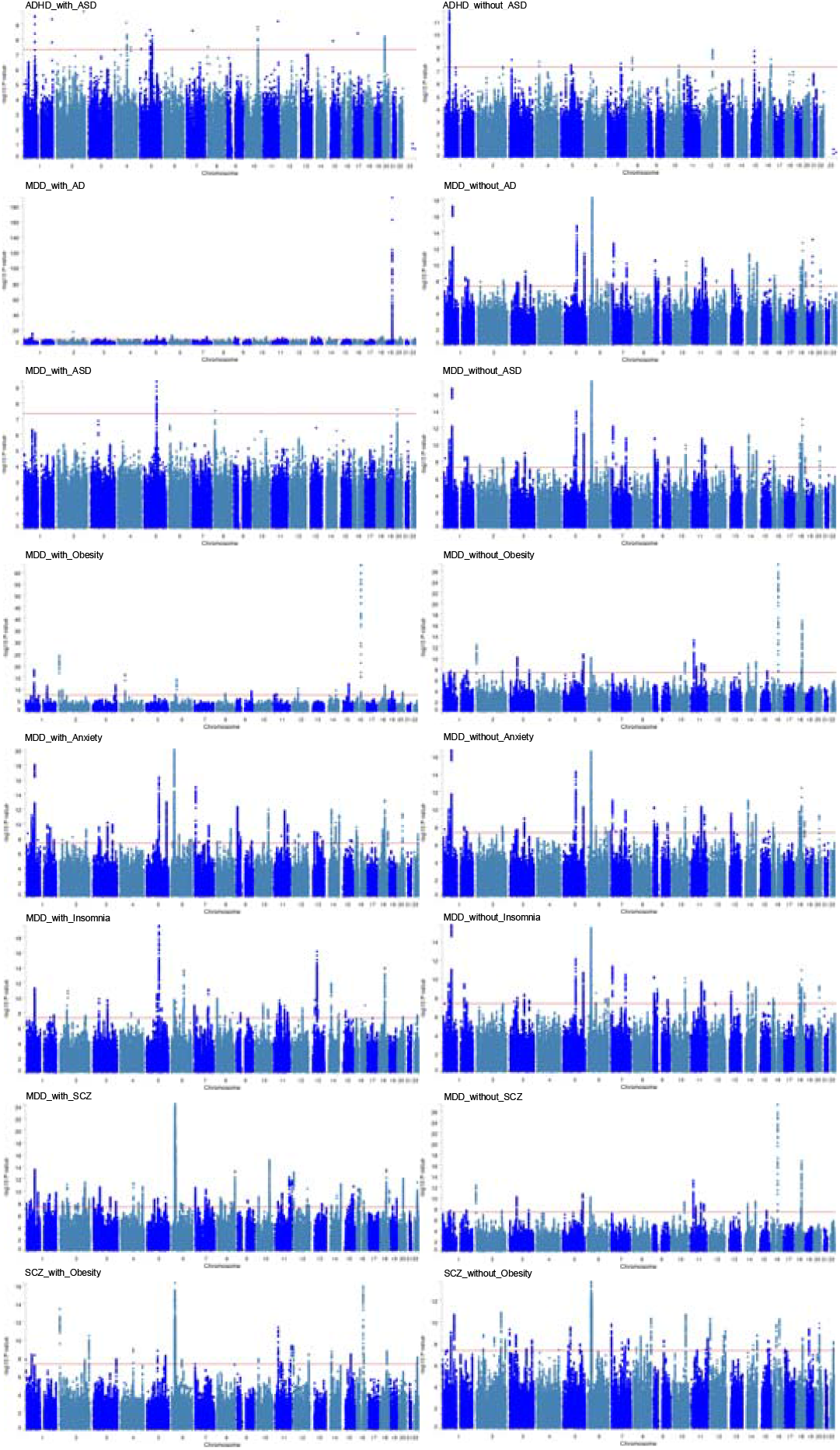
Manhattan plots of GWAS results for 16 psychiatric entities. Note: ADHD is the abbreviation for attention deficit hyperactivity disorder, ASD is the abbreviation for autism spectrum disorder, MDD is the abbreviation for major depressive disorder, SCZ is the abbreviation for schizophrenia

The identified susceptibility genetic variants were mapped to genes by MAGMA. Our analysis revealed a mixture of susceptibility genes shared across multiple psychiatric subtypes, as well as subtype-specific genes. Supplementary Table S3 provides additional details of the shared and non-shared genes associated with the psychiatric entities studied.

#### Tissue enrichment analysis

We conducted a tissue enrichment analysis using the FUMA tool, leveraging a pre-computed list of differentially expressed genes (DEGs) across various tissues obtained from GTEx. The input consisted of significant genes identified in GWAS (via MAGMA), which were then tested for enrichment among these DEGs. We view this analysis as hypothesis-generating, as differential expression does not necessarily imply a causal role for the tissue.

Supplementary Figure S1 shows the tissue enrichment analysis results for all 16 studied psychiatric entities studied. We observed that all the entities showed enrichment predominantly in brain tissues^23,24^. Interestingly, basal ganglia emerged as the top-enriched tissue for all psychiatric entities involving ADHD or ASD. Abundant evidence supports its critical role in maintaining normal motor actions and cognitive functions ^16-18^. In a study by Curtin et al. ^19^, ADHD was associated with 2.4-fold increased risk of basal ganglia diseases.

Another intriguing finding is that the cerebellum was the top-enriched tissue for all disease combinations involving MDD. A growing body of research has demonstrated its involvement in impaired cognitive function and emotion dysregulation ^20-22^. Compared to other psychiatric entities involving MDD, the two entities involving obesity (i.e., MDD with obesity and MDD without obesity) exhibited a lower number of enriched brain tissues, with cerebellar hemisphere ranked as the top-enriched tissue.

#### Pathway enrichment analysis

We conducted pathway enrichment analysis using the web tool ConsensusPathDB (detailed results in Supplementary Table S4). We observed a mixture of shared and condition-specific enriched pathways across different psychiatric entities. For instance, “Signaling by Nuclear Receptors” was one of the top-enriched pathways shared between MDD with AD and MDD without AD. In a previous study, Fries et al.^23^ have highlighted the critical role of this pathway in coordinating organism’s response to stress, a major risk factor for MDD. Additionally, Mandrekar-Colucci et al.^24^identified nuclear receptors as a promising therapeutic target for AD, as they could regulate microglial activation and mitigate the brain’s inflammatory responses.

The cardiolipin biosynthesis pathway was specifically enriched in MDD with AD. ^34^Alterations in cardiolipin content, structure, and localization have been reported to be associated with impaired neurogenesis and neuronal dysfunction^25^, contributing to neurodegenerative diseases.

Notably, we did not find any common pathways between ADHD with and without ASD, as well as between MDD with and without ASD.

#### Cell type enrichment analysis

Cell type enrichment analyses were also performed to uncover the involved cell types for each studied disease combinations (see Figure 2). However, since not all cell types were available for analysis in FUMA, our results should be considered exploratory. We found that GABAergic neurons were the most enriched cell types in most psychiatric entities. Fogoca et al.^26^ have reported that decreased expression of GABAergic interneuron markers in the frontal cortex could result in depressive-like behaviors. Moreover, impaired GABAergic neurotransmission has been implicated in patients with depression, prompting the development of therapeutic strategies targeting this deficit ^27^. It is noteworthy that the top-enriched cell types for the studied psychiatric disease combinations were all located in brain tissues.

**Figure 2.**
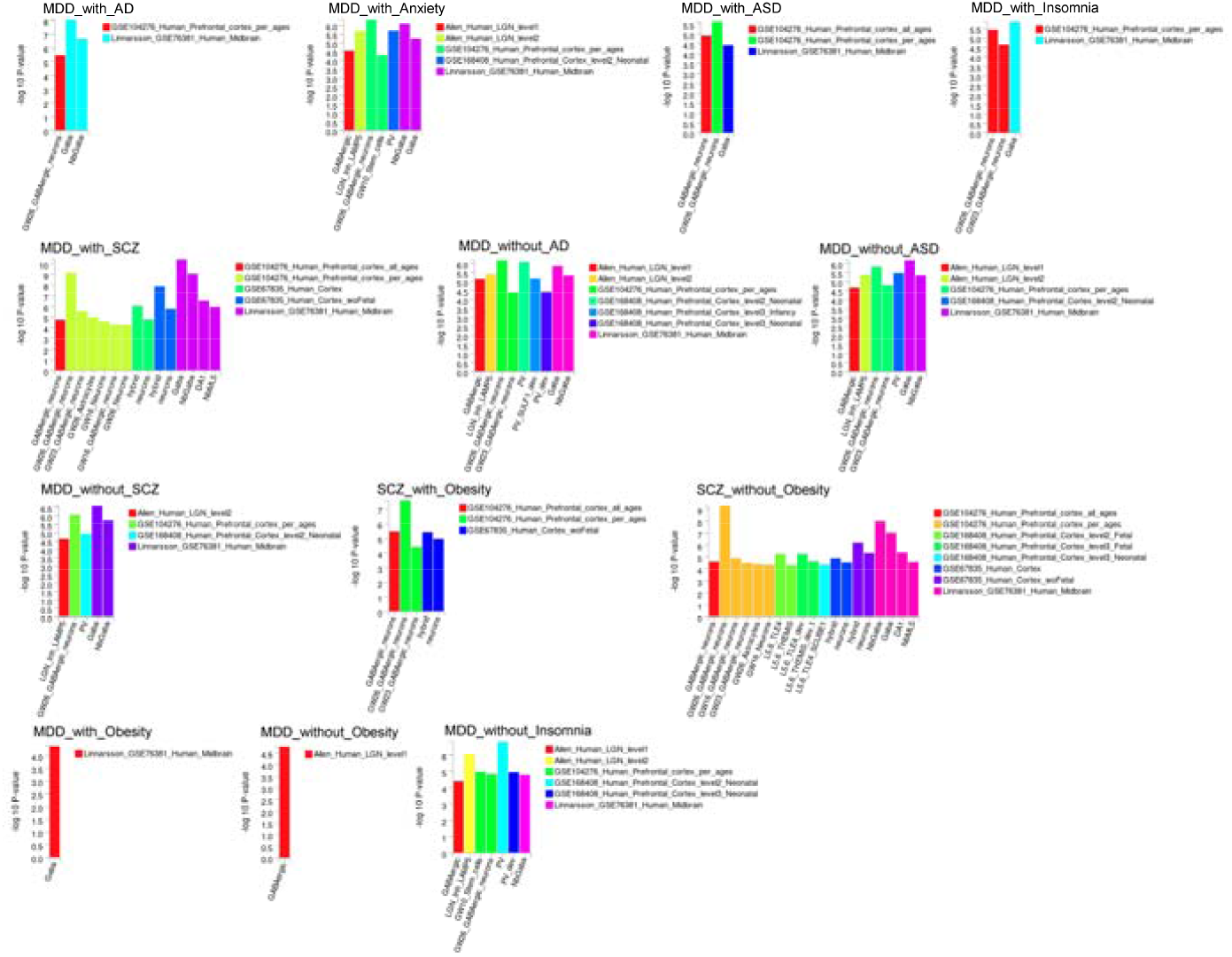
Cell type enrichment analysis results for each studied psychiatric disease combination

#### Drug enrichment analysis

To further explore the homogeneity and heterogeneity underlying the studied psychiatric entities, we performed drug enrichment analysis on the susceptibility gene sets identified for each disease combination (Supplementary Table S5). This analysis is considered exploratory or hypothesis-generating, as further experimental validations are required. Some enriched drugs were unique to particular psychiatric entities, while others were shared across different psychiatric entities. For example, chlorprothixene (a typical antipsychotic) was one of the top-enriched drugs specific to ADHD and ASD. Antipsychotics are often prescribed in ASD^28^ for irritability and associated behaviours including aggression and self-injury. Antipsychotics is also sometimes prescribed in ADHD due to other comorbid conditions, especially behaviour problems^29^. Among all disease combinations involved MDD, except for MDD with ASD, fendiline was among the top-enriched drugs. Interestingly, a previous study^30^ have suggested fendiline as a promising repurposing drug for MDD patients. For more detailed findings of drug enrichment analysis, please refer to Supplementary Table S5.

### Genetic correlation among psychiatric disease subtypes and cardiovascular diseases

We calculated genetic correlations between pairs of psychiatric entities with and without particular comorbid conditions (Table 2). The results revealed moderate to high genetic correlations in most pairs, suggesting shared genetic architecture. For instance, MDD with Anxiety Disorder exhibited a high genetic correlation with MDD without Anxiety Disorder (rg = 0.9964). Similarly, a high genetic correlation was also observed between SCZ with and without Obesity (rg = 0.9016). Interestingly, MDD with obesity only displayed a weak genetic correlation with MDD without obesity, implying they may represent *distinct biological subtypes* of MDD (rg = 0.1663).

**Table 2.** Genetic correlation between different psychiatric entities.

| 1st trait | 2nd trait | rg | P |
| --- | --- | --- | --- |
| MDD with Insomnia | MDD without Insomnia | 0.7915 (0.0117) | 0 |
| MDD with SCZ | MDD without SCZ | 0.5713 (0.0172) | 9.05E-243 |
| MDD with Obesity | MDD without Obesity | 0.1663 (0.0309) | 7.18E-08 |
| MDD with ASD | MDD without ASD | 0.6830 (0.0214) | 6.50E-224 |
| ADHD with ASD | ADHD without ASD | 0.8167 (0.0163) | <1E-245 |
| MDD with Anxiety | MDD without Anxiety | 0.9964 (0.001) | <1E-245 |
| MDD with AD | MDD without AD | 1.0255 (0.0265) | <1E-245 |
| SCZ with Obesity | SCZ without Obesity | 0.9016 (0.0133) | <1E-245 |

Here, rg indicates the calculated genetic correlation between two disease “subtypes”

To further investigate the genetic overlap between psychiatric entities and cardiovascular diseases, we analyzed their genetic correlations by LDSR (Table 3 and Table S6). As shown in Table 3, most “pairs” of psychiatric subtypes (i.e., a disorder with and without a comorbidity) exhibited significantly different genetic correlations with cardiovascular diseases, indicating distinct genetic relationships.

**Table 3.**
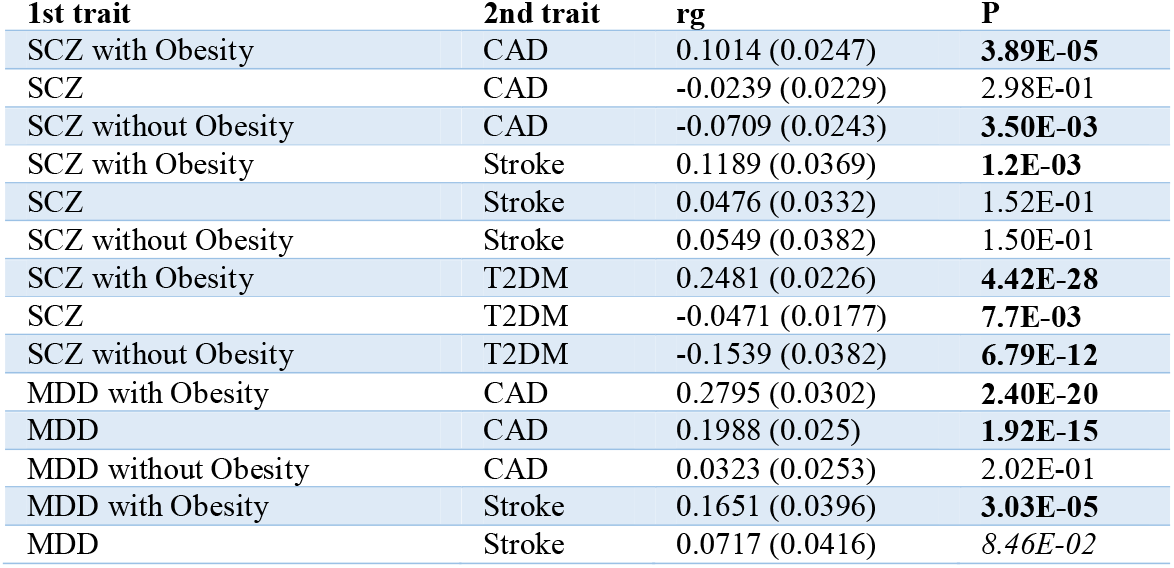
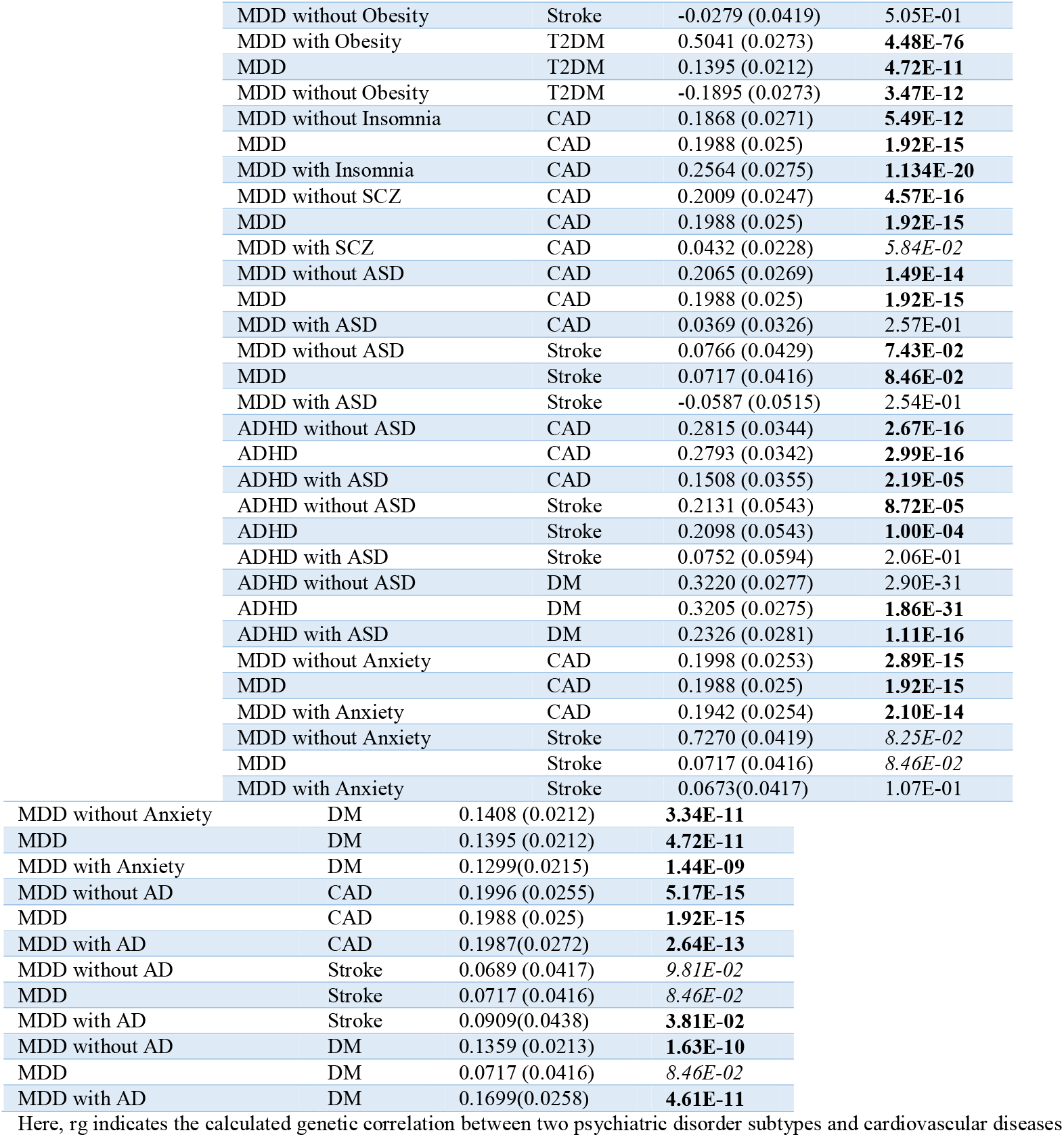
Genetic correlation between psychiatric entities and cardiovascular diseases.

| 1st trait | 2nd trait | rg | P |
| --- | --- | --- | --- |
| SCZ with Obesity | CAD | 0.1014 (0.0247) | **3.89E-05** |
| SCZ | CAD | -0.0239 (0.0229) | 2.98E-01 |
| SCZ without Obesity | CAD | -0.0709 (0.0243) | **3.50E-03** |
| SCZ with Obesity | Stroke | 0.1189 (0.0369) | **1.2E-03** |
| SCZ | Stroke | 0.0476 (0.0332) | 1.52E-01 |
| SCZ without Obesity | Stroke | 0.0549 (0.0382) | 1.50E-01 |
| SCZ with Obesity | T2DM | 0.2481 (0.0226) | **4.42E-28** |
| SCZ | T2DM | -0.0471 (0.0177) | **7.7E-03** |
| SCZ without Obesity | T2DM | -0.1539 (0.0382) | **6.79E-12** |
| MDD with Obesity | CAD | 0.2795 (0.0302) | **2.40E-20** |
| MDD | CAD | 0.1988 (0.025) | **1.92E-15** |
| MDD without Obesity | CAD | 0.0323 (0.0253) | 2.02E-01 |
| MDD with Obesity | Stroke | 0.1651 (0.0396) | **3.03E-05** |
| MDD | Stroke | 0.0717 (0.0416) | 8.46E-02 |
| MDD without Obesity | Stroke | -0.0279 (0.0419) | 5.05E-01 |
| MDD with Obesity | T2DM | 0.5041 (0.0273) | **4.48E-76** |
| MDD | T2DM | 0.1395 (0.0212) | **4.72E-11** |
| MDD without Obesity | T2DM | -0.1895 (0.0273) | **3.47E-12** |
| MDD without Insomnia | CAD | 0.1868 (0.0271) | **5.49E-12** |
| MDD | CAD | 0.1988 (0.025) | **1.92E-15** |
| MDD with Insomnia | CAD | 0.2564 (0.0275) | **1.134E-20** |
| MDD without SCZ | CAD | 0.2009 (0.0247) | **4.57E-16** |
| MDD | CAD | 0.1988 (0.025) | **1.92E-15** |
| MDD with SCZ | CAD | 0.0432 (0.0228) | 5.84E-02 |
| MDD without ASD | CAD | 0.2065 (0.0269) | **1.49E-14** |
| MDD | CAD | 0.1988 (0.025) | **1.92E-15** |
| MDD with ASD | CAD | 0.0369 (0.0326) | 2.57E-01 |
| MDD without ASD | Stroke | 0.0766 (0.0429) | **7.43E-02** |
| MDD | Stroke | 0.0717 (0.0416) | **8.46E-02** |
| MDD with ASD | Stroke | -0.0587 (0.0515) | 2.54E-01 |
| ADHD without ASD | CAD | 0.2815 (0.0344) | **2.67E-16** |
| ADHD | CAD | 0.2793 (0.0342) | **2.99E-16** |
| ADHD with ASD | CAD | 0.1508 (0.0355) | **2.19E-05** |
| ADHD without ASD | Stroke | 0.2131 (0.0543) | **8.72E-05** |
| ADHD | Stroke | 0.2098 (0.0543) | **1.00E-04** |
| ADHD with ASD | Stroke | 0.0752 (0.0594) | 2.06E-01 |
| ADHD without ASD | DM | 0.3220 (0.0277) | 2.90E-31 |
| ADHD | DM | 0.3205 (0.0275) | **1.86E-31** |
| ADHD with ASD | DM | 0.2326 (0.0281) | **1.11E-16** |
| MDD without Anxiety | CAD | 0.1998 (0.0253) | **2.89E-15** |
| MDD | CAD | 0.1988 (0.025) | **1.92E-15** |
| MDD with Anxiety | CAD | 0.1942 (0.0254) | **2.10E-14** |
| MDD without Anxiety | Stroke | 0.7270 (0.0419) | 8.25E-02 |
| MDD | Stroke | 0.0717 (0.0416) | 8.46E-02 |
| MDD with Anxiety | Stroke | 0.0673 (0.0417) | 1.07E-01 |
| MDD without Anxiety | DM | 0.1408 (0.0212) | **3.34E-11** |
| MDD | DM | 0.1395 (0.0212) | **4.72E-11** |
| MDD with Anxiety | DM | 0.1299 (0.0215) | **1.44E-09** |
| MDD without AD | CAD | 0.1996 (0.0255) | **5.17E-15** |
| MDD | CAD | 0.1988 (0.025) | **1.92E-15** |
| MDD with AD | CAD | 0.1987 (0.0272) | **2.64E-13** |
| MDD without AD | Stroke | 0.0689 (0.0417) | 9.81E-02 |
| MDD | Stroke | 0.0717 (0.0416) | 8.46E-02 |
| MDD with AD | Stroke | 0.0909 (0.0438) | **3.81E-02** |
| MDD without AD | DM | 0.1359 (0.0213) | **1.63E-10** |
| MDD | DM | 0.0717 (0.0416) | 8.46E-02 |
| MDD with AD | DM | 0.1699 (0.0258) | **4.61E-11** |

Here, rg indicates the calculated genetic correlation between two psychiatric disorder subtypes and cardiovascular diseases

Interestingly, some psychiatric subtypes displayed opposite genetic correlations with the same cardiovascular disease. For example, while SCZ with Obesity exhibited a positive genetic correlation with type 2 diabetes (T2DM) (rg=0.2481), SCZ without Obesity demonstrated a negative genetic correlation with T2DM (rg= -0.1539). Similarly, SCZ with and without obesity displayed opposing genetic correlations with coronary artery disease (CAD). Notably, the genetic correlations between “un-subtyped”(originally defined) psychiatric disorders and cardiovascular diseases generally lie between those for “subtyped”(comorbid or only single) disorders. These findings highlight the complex interactions between genetic factors associated with psychiatric entities and cardiovascular diseases, indicating potential distinct genetic signatures for different psychiatric subtypes.

### Results of MR analysis

MR analysis was conducted to investigate whether a certain psychiatric disease combination is causally linked to a significantly higher or lower risk of cardiovascular outcomes. Notably, most results were consistent across different *r*^2^ cutoffs. For simplicity, we primarily report analysis results at *r*^2^ = 0.001, *p*= 5E - 08, which are standard settings in TwoSampleMR. Table 4 summarizes the results (please refer to Supplementary Table S7 for full results). Note that with a binary exposure, the causal estimate (odds ratio, OR) reflects the average effect on the outcome associated with a 2.72-fold increase in the prevalence of the exposure (e.g., an increase in exposure prevalence from 1% to 2.72%)^31^.

**Table 4.** MR analysis results for cardiovascular outcomes.

| Exposure | Outcome | OR(2.72 times of exposure preval) | P | Exposure | Outcome | OR(2.72 times of exposure preval) | P |
| --- | --- | --- | --- | --- | --- | --- | --- |
| ADHD with ASD | CAD | 1.350 | 2.04E-01 | ADHD without ASD | CAD | 1.047 | 2.71E-01 |
| ADHD with ASD | DM | 0.312 | **8.05E-02\*** | ADHD without ASD | DM | 1.102 | **1.58E-05** |
| ADHD with ASD | Stroke | 1.680 | **4.84E-02** | ADHD without ASD | Stroke | 1.083 | **4.76E-02** |
| MDD with AD | CAD | 1.310 | **1.97E-10** | MDD without AD | CAD | 0.594 | 1.1.3E-01* |
| MDD with AD | DM | 0.712 | **2.14E-05\*** | MDD without AD | DM | 1.224 | **4.23E-03** |
| MDD with AD | Stroke | 1.059 | 3.87E-01 | MDD without AD | Stroke | 1.037 | 5.36E-01 |
| MDD with Anxiety | CAD | 2.106 | **2.35E-05** | MDD without Anxiety | CAD | 1.108 | **3.78E-05** |
| MDD with Anxiety | DM | 1.917 | **3.03E-03** | MDD without Anxiety | DM | 1.109 | **3.76E-04** |
| MDD with Anxiety | Stroke | 1.537 | **5.79E-02** | MDD without Anxiety | Stroke | 1.045 | 1.56E-01 |
| MDD with ASD | CAD | 1.088 | 1.69E-01 | MDD without ASD | CAD | 1.184 | **3.60E-05** |
| MDD with ASD | DM | 1.007 | 9.82E-01 | MDD without ASD | DM | 1.166 | **1.99E-03** |
| MDD with ASD | Stroke | 1.062 | 5.90E-01 | MDD without ASD | Stroke | 1.058 | 2.70E-01 |
| MDD with Insomnia | CAD | 3.721 | **1.72E-06** | MDD without Insomnia | CAD | 1.054 | **2.71E-03** |
| MDD with Insomnia | DM | 2.115 | **1.59E-02** | MDD without Insomnia | DM | 1.056 | **7.31E-03** |
| MDD with Insomnia | Stroke | 1.340 | 2.67E-01 | MDD without Insomnia | Stroke | 1.027 | 2.46E-01 |
| MDD with Obesity | CAD | 1.365 | **1.81E-07** | MDD without Obesity | CAD | 0.919 | 1.94E-01 |
| MDD with Obesity | DM | 5.418 | **3.58E-06\*** | MDD without Obesity | DM | 0.292 | **9.87E-04\*** |
| MDD with Obesity | Stroke | 1.078 | 2.09E-01 | MDD without Obesity | Stroke | 0.943 | **8.52E-02** |
| MDD with SCZ | CAD | 1.059 | 2.28E-01 | MDD without SCZ | CAD | 1.211 | **4.64E-05** |
| MDD with SCZ | DM | 0.964 | 3.63E-01 | MDD without SCZ | DM | 1.182 | **3.52E-03** |
| MDD with SCZ | Stroke | 1.031 | 4.92E-01 | MDD without SCZ | Stroke | 1.061 | 3.28E-01 |
| SCZ with Obesity | CAD | 1.117 | **1.83E-02** | SCZ without Obesity | CAD | 1.156 | **2.91E-02\*** |
| SCZ with Obesity | DM | 1.364 | **2.81E-03** | SCZ without Obesity | DM | 0.948 | **2.58E-02** |
| SCZ with Obesity | Stroke | 1.106 | **6.94E-02** | SCZ without Obesity | Stroke | 1.009 | 5.43E-01 |

Note: MR analysis results were primarily based on MR-IVW; \* indicates that the causal analysis results were derived from MR-Egger instead of MR-IVW due to the presence of imbalanced horizontal pleiotropy, as indicated by a significant Egger intercept. Since the exposure is binary, the OR represents the average effect to the outcome if the exposure prevalence (preval) is increased by 2.72 times.

Many studied psychiatric disorder combinations exhibited a significant causal relationship with increased risks of cardiovascular diseases. Importantly, the magnitude of effect sizes varies across different psychiatric disorder subtypes. For example, both MDD with and without Anxiety were found to be causally linked to significantly increased CAD risks. The OR of MDD with Anxiety on CAD is 2.106 (MR-IVW, p=2.35E-05), while that of MDD without Anxiety on CAD is 1.108 (MR-Egger, p=3.78E-05).

Additionally, we found that comorbid psychiatric disorders often represent more substantial risk factors for cardiovascular outcomes than single disorders. For example, MDD with Insomnia was causally linked to a significantly increased risk of CAD with an OR of 3.721 (MR-IVW, p=1.72E-06), while MDD without Insomnia had a much lower OR of 1.054 (MR-IVW, p=2.71E-03).

Both obese MDD and obese SCZ were found to be causally linked to increased T2DM risk. Specifically, obese MDD and obese SCZ were linked to T2DM risk with ORs of 5.418 (MR-IVW, p = 3.58E-06) and 1.364 (MR-Egger, p = 2.81E-03), respectively. In contrast, non-obese MDD (MR-Egger, OR=0.292, p=9.87E-04) and non-obese SCZ (MR-Egger, OR=0.948, p=2.58E-02) were associated with significantly *decreased* T2DM risk.

The above results underscore the complex causal relationships between specific psychiatric disease combinations and various cardiovascular outcomes. ^43^

## Discussion

### Overview

This study applied the CombGWAS statistical framework to uncover the genetic basis underlying different combinations of psychiatric disorders. Specifically, we studied the genetic architecture of 16 distinct psychiatric entities, comprising 8 pairs of subtypes representing comorbid and single psychiatric disorders, respectively. We successfully identified a total of 653 susceptibility genomic risk loci associated with these psychiatric disorder entities. Among these loci, 325 were found to be specifically associated with comorbid disorders, while 328 were associated with single disorders.

We conducted comprehensive secondary analyses to examine shared and distinct genetic signatures across the 8 pairs of psychiatric subtypes. These analyses provided insights into the genetic factors contributing to the development and manifestation of these specific subtypes.

Here we also highlight a few top genes identified for several psychiatric disorder subtypes. For instance, *MANBA* and *XRN2* emerged as the top susceptibility genes identified in both ADHD with and without ASD. This finding aligns with previous research by Peyre et al ^32^, who reported these two genes as susceptibility genes shared by ADHD and ASD.

Notably, *SORCS3* was found to be associated with all 12 studied psychiatric entities involving MDD. Previous studies have highlighted its critical role in regulating neuronal viability and function ^33-35^.

Furthermore, Brediderhoff et al. ^36^ suggested that *SORCS3* may induce damage to fear memory modulation through altered synaptic plasticity.

### Shared and unique genetic underpinnings of psychiatric disorder subtypes

Shared susceptibility genes were identified across all 8 pairs of subtypes, suggesting an overlap of genetic risks across different combinations of psychiatric disorders. Notably, the proportion of shared genes varies across different subtype pairs (Table 5). Specifically, we observed substantial overlap of susceptibility genes between MDD with Anxiety Disorder and MDD without Anxiety Disorder. Conversely, only a small proportion of shared genes were found between MDD with Obesity and MDD without Obesity. These shared genes provide insights into the biological similarities between subtypes and potential shared pathophysiology. Additionally, the identification of subtype-specific susceptibility genes could be utilized in the differentiation between different subtypes and inform targeted treatment strategies.

**Table 5.** Number of shared genomic risk loci, genes and pathways across 8 pairs of subtypes.

| 1st trait | 2nd trait | Loci |  |  | Genes |  |  | Pathways |  |  |
| --- | --- | --- | --- | --- | --- | --- | --- | --- | --- | --- |
|  |  | 1st trait only | 2nd trait only | shared | 1st trait only | 2nd trait only | shared | 1st trait only | 2nd trait only | shared |
| MDD with Insomnia | MDD without Insomnia | 46 | 37 | 2 | 517 | 307 | 263 | 25 | 22 | 81 |
| MDD with SCZ | MDD without SCZ | 95 | 52 | 0 | 2054 | 337 | 423 | 48 | 13 | 91 |
| MDD with Obesity | MDD without Obesity | 21 | 21 | 0 | 126 | 271 | 23 | 6 | 87 | 0 |
| MDD with ASD | MDD without ASD | 3 | 54 | 0 | 25 | 744 | 19 | 0 | 104 | 0 |
| ADHD with ASD | ADHD without ASD | 18 | 11 | 1 | 22 | 49 | 20 | 1 | 9 | 0 |
| MDD with Anxiety | MDD without Anxiety | 48 | 27 | 25 | 346 | 63 | 643 | 6 | 9 | 97 |
| MDD with AD | MDD without AD | 31 | 47 | 6 | 189 | 411 | 307 | 26 | 3 | 98 |
| SCZ with Obesity | SCZ without Obesity | 29 | 48 | 0 | 320 | 1239 | 517 | 9 | 21 | 100 |

Furthermore, we performed comprehensive analysis of enriched pathways, cell types, and drugs based on the GWAS results. These enrichment analyses shed light on the potential biological mechanisms and pathways involved in the development and manifestation of specific subtypes. We also revealed some interesting patterns. For example, although only a moderate proportion of shared susceptibility genes was observed between SCZ with Obesity and SCZ without Obesity, the number of shared pathways significantly exceeds the number of unique pathways (Table 5). These findings suggest that the proportion of shared susceptibility genes does not necessarily correspond to the proportion of shared pathways between subtypes. For example, it can possibly be due to the fact that different genes may function in the same pathways.

### Genetic correlations among disease subtypes and cardiovascular diseases

The majority of disease subtypes exhibited high (rg>=0.8) or moderate (0.4=<rg<0.8) correlation with one another. Among the 8 pairs of subtypes examined, only one pair, i.e., MDD with Obesity vs MDD without Obesity, demonstrates a weak correlation. Despite their high genetic correlations, most subtypes exhibited distinct genetic correlations with the same cardiovascular disease. Some pairs even displayed entirely opposing genetic correlations, particularly those subtype pairs that involved obesity. These findings suggest that the psychiatric disorder combinations we studied may represent biologically distinct subtypes with both shared and unique genetic signatures.

Evidence indicates that normal weight may protect against cardiovascular diseases^37^, potentially counteracting the elevated risks of CVD conferred by psychiatric disorders. This aligns with the observed opposing or weak genetic correlations between non-obese psychiatric disorders and CVD.

### Varying causal effects of different psychiatric disorder subtypes on cardiovascular outcomes

While many psychiatric disorder combinations showed increased cardiovascular disease risks, comorbid disorders typically demonstrated stronger causal effects than single disorders. This highlights the importance of considering the cumulative impact of comorbid disorders on cardiovascular health.

Of note, obese and non-obese MDD/SCZ showed opposite causal effects on T2DM risk. These findings underscore the need for more comprehensive and personalized screening, prevention, and treatment strategies that address both psychiatric and cardiovascular conditions in individuals with comorbidities.

### Limitations

This study has several limitations. Firstly, the GWAS summary statistics used were primarily derived from European samples. As such, some results may be population specific and may not be generalizable to non-European groups. This limitation may be mitigated with the availability of summary statistics from other ethnic groups. While our analysis provides valuable insights into the genetic signatures underlying different psychiatric disease subtypes, further exploration is required to validate these findings. Replication studies using independent cohorts and individuals with comorbid conditions, as well as more diverse populations would enhance the robustness and generalizability of the observed genetic associations. Besides, our study only included a limited number of psychiatric disease combinations. Also, the biological mechanisms underlying the psychiatric disorder subtypes will require further experimental studies.

## Conclusions

To conclude, our study successfully identified the genetic basis and risk genes for 8 pairs of psychiatric disease subtypes. Secondary analyses, including pathway, cell type, and drug enrichment analyses, provided further biological insights into the pathophysiology of these disorder subtypes. Genetic correlation and MR analyses shed light on the links of these subtypes with various CVD, which may have important clinical implications. Furthermore, the identified non-shared susceptibility genes may provide a way to differentiate the different ‘subtypes’.

Future research should incorporate data from diverse ethnic groups, conduct replication studies, and perform functional investigations to enhance the robustness and generalizability of the observed genetic associations. By elucidating the genetic basis of a wide variety of psychiatric disorder subtypes, our study paves the way for the development of personalized treatment and preventive strategies for individuals affected by these complex disorders.

## Data Availability

All GWAS summary statistics used in this study were publicly available at PGC, iPSYCH,CTG and GIANT consortium

## Data availability

The GWAS summary statistics of studied disorders were downloaded from publicly available database.

## Acknowledgements

This work was supported partially by a National Natural Science Foundation China (NSFC) Young Scientist Grant (31900495), a National Natural Science Foundation China grant (81971706), the Lo Kwee Seong Biomedical Research Fund from The Chinese University of Hong Kong and the KIZ-CUHK Joint Laboratory of Bioresources and Molecular Research of Common Diseases, Kunming Institute of Zoology and The Chinese University of Hong Kong, China.

