## Supplementary table file for "Uncovering the genetic underpinnings for different psychiatric disorder combinations"

Table S1 Summary of GWAS data for 8 psychiatric (related) disorders

| **Traits/Disorders** | **Abbreviation** | **Source** | **Data type** | **Total N** | **Prevalence (%)** |
| --- | --- | --- | --- | --- | --- |
| Major Depression Disorder (2018) | MDD | PGC | binary | 173,005 | 13.0 ^1^ |
| Schizophrenia (2018) | SCZ | Pardiñas et al. | binary | 320,404 | 0.5 ^2^ |
| Autism Spectrum Disorder (2019) | ASD | iPSYCH&PGC | binary | 46,350 | 2.5 ^3^ |
| Attention deficit hyperactivity disorder (2019) | ADHD | iPSYCH&PGC | binary | 53,293 | 6.5 ^4^ |
| Alzheimer’s disease | AD | CTG lab | binary | 455,258 | 10.7 ^5^ |
| Obesity | Obesity | GIANT consortium | binary | 40585 | 30 ^6^ |
| Anxiety Disorder (2018) | Anxiety | UKBB | binary | 117,751 | 14.2 ^7^ |
| Insomnia (2019) | Insomnia | UKBB&CTG lab | binary | 386,533 | 10.0 ^8^ |

Table S2 Identified genomic risk loci for different psychiatric disease combinations

Table S3 Identified susceptibility genes for each studied psychiatric disease combination

Table S4 Enriched pathways for 16 studied psychiatric disease combinations

Table S5 Drug enrichment analysis based on identified susceptibility genes for studied disease combinations

Table S6 MR analysis results for cardiovascular outcomes by MR Egger and inverse variance weighted at multiple pvalue thresholds
