## Supplementary figures for "Uncovering the genetic underpinnings for different psychiatric disorder combinations"

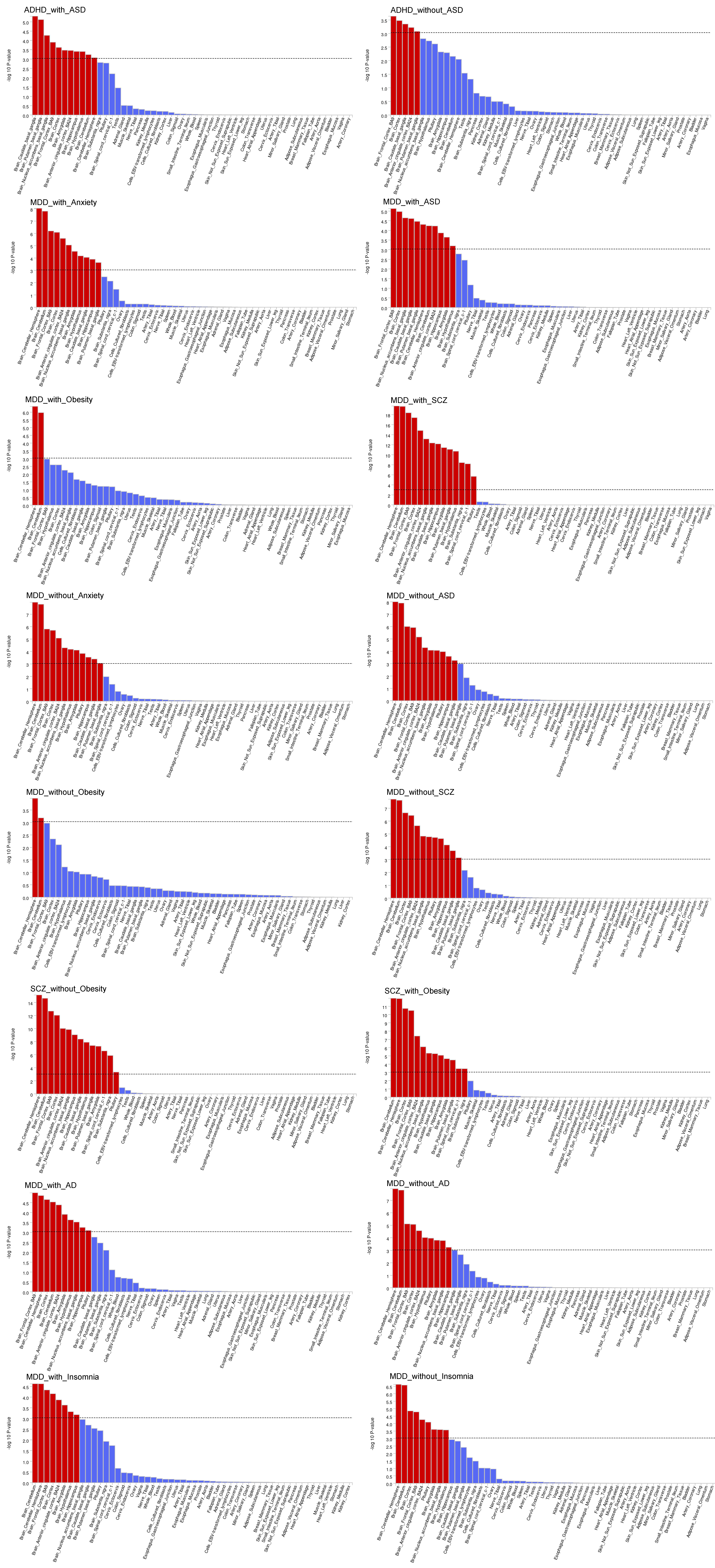

Fig. S1 Tissue specificity analysis results for each studied psychiatric disease combination
